# A Nationally Representative Study of Complementary and Alternative Medicine Therapies in relation to Sleep Duration and Insomnia Symptoms among US Adults

**DOI:** 10.64898/2026.08.17.26360473

**Authors:** Rupsha Singh, Symielle A. Gaston, Christopher Payne, Dayna T. Neo, Suzanne M. Bertisch, Chandra L. Jackson

## Abstract

**Background:** Complementary and Alternative Medicine (CAM) therapies, such as massage, meditation, and yoga, are widely used to promote wellness, including sleep improvement. Although some CAM therapies may improve sleep through stress reduction, relaxation, and relief of physical discomfort, little is known about associations between individual CAM modalities and sleep health at the population level. Therefore, we investigated the associations between CAM therapies and short sleep duration as well as insomnia symptoms.

**Methods:** Participants from the nationally-representative 2012 National Health Interview Survey (NHIS) self-reported the use of CAM therapies and short sleep duration (<7 hours vs. 7–9 hours) as well as insomnia symptoms (yes vs. no). Poisson regression with robust variance was used to estimate adjusted prevalence ratios (aPRs) and 95% confidence intervals (CIs) for cross-sectional associations between CAM therapy use and sleep outcomes.

**Results:** Among 30,405 participants, the average age was 46.1 ± 0.2 years and 51% were women. Adults reporting any vs. no CAM therapy use had a higher prevalence of short sleep duration (aPR: 1.11; 95% CI: 1.05–1.16) and insomnia symptoms (aPR: 1.56; 95% CI: 1.47– 1.65) after adjustment for sociodemographic and clinical characteristics. Herbal supplements (aPR: 1.13; 95% CI: 1.07–1.19) and massage (aPR: 1.14; 95% CI: 1.05–1.23) were associated with higher prevalence of short sleep duration. Most CAM therapies were associated with higher prevalence of insomnia symptoms, with the strongest associations observed for meditation/guided imagery/progressive relaxation (aPR: 1.84; 95% CI: 1.68–2.02).

**Conclusion:** The higher prevalence of short sleep duration and insomnia symptoms among CAM users may reflect reverse causation, as adults with more severe or persistent sleep disturbances may be more likely to seek CAM therapies. Longitudinal studies are needed to clarify directionality.

## INTRODUCTION

Sleep is an essential pillar of physical and mental health. Sleep disturbances and sleep disorders have been associated with numerous adverse health outcomes, including obesity, hypertension, type 2 diabetes, cardiovascular disease, depression, anxiety, impaired cognitive functioning, reduced quality of life, and premature mortality [1–9]. Despite the well-established importance of sleep health, sleep disturbances remain highly prevalent in the United States. Approximately, 33% of US adults reported short sleep duration and approximately 50 to 70 million Americans have a sleep disorder [10, 11]. Insomnia symptoms, including difficulty initiating sleep, difficulty maintaining sleep, and early morning awakenings, are among the most common sleep complaints and are associated with impaired daytime functioning, reduced emotional well-being, poorer quality of life, and increased burden of co-occurring conditions such as stress, anxiety, depression, chronic pain, and cardiometabolic disease [6, 12, 13].

Although cognitive behavioral therapy for insomnia – considered the gold standard – and pharmacologic treatments are commonly used for insomnia symptoms and sleep disorders, multiple barriers may limit access to evidence-based sleep care. These barriers include limited availability of behavioral sleep medicine providers, cost and accessibility concerns, long wait times, stigma surrounding mental health treatment, concerns regarding medication dependence, side effects, and long-term pharmacologic use, and patient preference [14–16]. Consequently, many adults may self-treat or seek using over-the-counter or alternative treatments for sleep disorders or sleep disturbances more broadly. Complementary and Alternative Medicine (CAM) therapies refer to alternative or adjunctive approaches that are commonly used for improving sleep, stress, emotional well-being, and overall health [17, 18]. CAM refers to a broad range of health practices, therapies, and medical systems that are not traditionally considered part of conventional Western medicine and are often used to promote holistic healing, symptom management, and overall well-being [19]. NIH classifications of these therapies include manipulative and body-based practices (e.g., reflexology and massage therapy), mind-body therapies (e.g., yoga and meditation), biologically-based practices (e.g., herbal and dietary supplements), energy-based practices (e.g., reiki), and lastly, medical systems rooted in non-Western traditions (e.g., Ayurvedic medicine and traditional Chinese Medicine) [20]. Although this field of practice is not considered standard medical care, many adults report using these practices alongside (‘complementary’) or in lieu of (‘alternative’) conventional medicine [21, 22]. In the United States, CAM utilization has increased substantially over recent decades, with 2007 National Health Interview Survey (NHIS) data indicating that approximately 40% of US adults reported using at least one CAM therapy within the previous 12 months [23]. Increasing use of CAM therapies has been attributed to multiple factors, including growing interest in holistic and whole-person approaches to health, concerns regarding medication side effects, dissatisfaction with conventional treatments, preferences for natural remedies, cultural beliefs and practices, and greater emphasis on self-management and wellness-oriented healthcare approaches [18, 24]. Recent perspectives on whole-person sleep health further emphasize that sleep is influenced by interconnected biological, psychological, social, cultural, and spiritual dimensions of health, reinforcing interest in integrative approaches to sleep care [25]. Emerging epidemiologic evidence further suggests that religion and spirituality may be associated with sleep health, particularly in the context of psychosocial stress, underscoring the value of multidisciplinary sleep care [26]. Additionally, culturally rooted healing traditions often conceptualize sleep within this broader context, which may further contribute to CAM utilization among adults experiencing sleep difficulties [27–29].

Recent studies have shown that adults frequently report the use of CAM therapies for insomnia, sleep apnea, and sleep disturbances [30–35]. Prior research has largely focused on whether specific CAM interventions improve subjective sleep quality, insomnia severity, sleep latency, or related symptoms, particularly mindfulness-based movement and mind-body therapies such as yoga, meditation, Tai Chi, Qigong, acupuncture, acupressure, and relaxation techniques [30, 33, 34, 36, 37]. Several systematic reviews and meta-analyses suggest that some CAM approaches, particularly mind-body interventions, may improve subjective sleep quality and insomnia symptoms, although evidence quality remains heterogeneous or mixed across intervention types and study populations [30, 37, 38]. For example, a systematic review and meta-analysis of 18 randomized controlled trials found that mindfulness meditation improved sleep quality compared with nonspecific active controls, although it was not superior to evidence-based sleep treatments [22]. Mindfulness-based practices, including yoga, Tai Chi, Qigong, and meditation, have also been associated with improved sleep quality and reduced insomnia symptoms [33, 35, 36]. At the same time, population-based studies suggest that adults with insomnia symptoms are more likely to use CAM than those without insomnia, likely reflecting treatment-seeking behavior rather than necessarily better sleep among CAM users [17]. Similarly, among patients with obstructive sleep apnea, 58% reported ever using a CAM therapy, 21% reported current use, and 58% expressed interest in future CAM use to improve sleep [31]. These findings suggest a complex relationship between CAM use and sleep health, in which CAM use may reflect both attempts to manage existing sleep problems and engagement in health-promoting behaviors. However, despite growing evidence regarding CAM therapies and insomnia-related outcomes, limited research has examined associations between individual CAM therapies and sleep health at the population level. Therefore, the purpose of this study was to examine cross-sectional associations between individual CAM therapies and both short sleep duration and insomnia symptoms among US adults using nationally representative NHIS data. Given evidence that some CAM therapies may improve sleep, but that individuals with sleep problems are also more likely to seek CAM therapies, we hypothesized that CAM therapy use, compared to no use, is associated with short sleep duration and insomnia symptoms.

## METHODS

### Data Source

Using the Integrated Public Use Microdata (IPUMS) [39], we analyzed cross-sectional data from the 2012 National Health Interview Survey (NHIS). The NHIS is an annual household survey of the non-institutionalized, US population that uses a multistage sampling design to obtain a nationally representative sample. Beginning in 2002 and ending in 2012, every five years the NHIS included an alternative health questionnaire on CAM use in addition to the core questionnaire. The 2012 NHIS consisted of 34,525 sample adults, all of whom provided informed consent, with a conditional response rate of 79.7% [40]. Analysis of de-identified, secondary data is not deemed human subjects research; therefore, approval by the National Institutes of Health Institutional Review Board was not required.

### Study population

Included in this complete-case analysis were participants aged 18 years and older (n=34,525). Excluded from this analysis were participants missing information for usual sleep duration (n=493), insomnia or trouble sleeping (n=5), CAM therapies (n=914), race and ethnicity (n=66), educational attainment (n=122), household income (n=1,683), employment status (n=27), marital status (n=37), body mass index (n=764), or general health status (n=9). The final analytic sample consisted of 30,405 participants (Supplemental Figure 1). Compared with included participants, excluded participants were generally older and more likely to be women, have lower educational attainment and household income, be unemployed or not in the labor force, and report poorer health behaviors and overall health (Supplemental Table 1).

### Exposure Assessment: Complementary and Alternative Medicine Therapies

The 2012 NHIS adult alternative core questionnaire asked participants a series of questions about their use of CAM therapies. These CAM therapies included acupuncture, ayurveda, biofeedback, chelation therapy, chiropractic or osteopathic manipulation, craniosacral therapy, energy healing therapy, herbal and non-vitamin supplements, homeopathy, hypnosis, massage, meditation, guided imagery, and progressive relaxation techniques, movement and exercise techniques, naturopathy, special diets, traditional healers, and yoga/tai chi/qigong (see Appendix I for more information).

Traditional healers included Native American Healers/Medicine Men, Shaman, Curandero/Machi/Parchero, Yerbero/Hierbista, Sobador, or Huesero (see Appendix II for more information). Herbal and non-vitamin supplements consisted of 20 supplements that included pills, capsules, tablets, or liquids commonly labeled as dietary supplements but did not include vitamin or mineral supplements, homeopathic treatments, or herbal/green teas (see Appendix III for the complete list). Meditation, guided imagery, and progressive relaxation techniques consisted of mantra meditation including transcendental meditation, relaxation response, and clinically standardized meditation, mindfulness meditation including Vipassana, Zen Buddhist meditation, mindfulness-based stress reduction, and mindfulness-based cognitive therapy, spiritual meditation including centering prayer and contemplative meditation, guided imagery, and progressive relaxation (see Appendix IV for more information). Movement and exercise techniques included Feldenkrais method, Alexander technique, Pilates, and Trager Psychophysical Integration (see Appendix V for more information). Special diets consisted of a vegetarian/vegan diet, a macrobiotic diet, The Atkins diet, The Pritikin diet, and The Ornish diet (see Appendix VI for more information).

We focused on the survey questions that asked respondents to describe their CAM therapy use. For most CAM therapies, participants were asked whether they had ever used the therapy or ever seen a practitioner of the therapy and if so, participants were asked whether they had done so in the past 12 months. Additional questions assessed the frequency of use, reasons for use, and associated costs. For the present analysis, CAM use was coded as a binary variable, with respondents classified as users (1) if they reported using the therapy and/or seeing a practitioner for the therapy within the past 12 months and non-users (0) if they reported never using the therapy, never seeing a practitioner, or not having used the therapy or seen a practitioner within the past 12 months. For CAM categories that included multiple modalities (e.g., traditional healers, mind-body relaxation therapies, mind-body exercise therapies, special diets, and movement or exercise techniques), respondents were classified as users if they reported use of at least one modality within that category during the past 12 months. Responses of “Don’t know” or “Refused” were coded as missing.

### Outcome Assessment: Sleep duration and insomnia symptoms

We analyzed the two sleep measures available for all study participants, self-reported short sleep duration and insomnia symptoms. Short sleep duration was derived from the question, “On average, how many hours of sleep do you get in a 24-hour period?” and dichotomized to short sleep duration (<7 hours) and recommended sleep duration (7-9 hours). Interviewers were instructed to round half hours to the nearest whole hour. Responses of “Don’t know” and “Refused” were combined and coded as missing. Long sleep duration (>9 hours) was not analyzed due to low prevalence. Data on insomnia symptoms (yes vs. no) was derived from the question, “During the past 12 months, have you…Regularly had insomnia or trouble sleeping?”. Responses of “Don’t know” and “Refused” were coded as missing.

### Potential confounders

Potential confounding variables were selected *a priori* based on prior literature and their potential associations with both CAM therapy use and sleep outcomes [41]. Sociodemographic characteristics included age (years), sex (male, female), race and ethnicity (Hispanic/Latine, Non-Hispanic (NH)-American Indian/Alaska Native, NH-Asian, NH-Black/African American, NH-Multiple race groups, NH-White), educational attainment (<high school, high school graduate or GED equivalent, some college including technical/vocational/occupational training or associate’s degree, ≥college including bachelor’s, master’s, professional, or doctoral degree), household income (<$35,000, $35,000–$74,999, ≥$75,000), employment status (employed full-time, employed part-time, unemployed), marital status (married/living with partner, divorced/separated/widowed, never married), and region of residence (Northeast, Midwest, South, West). Health-related characteristics included body mass index (underweight/recommended [<25 kg/m²], overweight [25–<30 kg/m²], obesity [≥30 kg/m²]) and self-rated health status (excellent, very good, good, fair/poor).

### Statistical Analysis

Descriptive statistics were estimated overall and by any CAM therapy use. We report survey-weighted means (± standard errors) for continuous variables and survey-weighted proportions for categorical variables. All analyses incorporated NHIS sample weights, strata, and primary sampling units using Stata survey estimation procedures to account for the complex multistage sampling design. The sampling weights represent the inverse probability of selection, adjusted for non-response with post-stratification adjustments. Poisson regression with robust standard errors was used to estimate prevalence ratios and 95% confidence intervals for associations between individual CAM therapy use and self-reported short sleep duration and insomnia adjusting for potential confounders. The significance level was set at a two-sided p-value of 0.05. All analyses were performed using Stata, Version 19.5 (Statacorp, College Station, Texas).

## RESULTS

### Study population characteristics

Among 30,405 participants, the average age was 46.1 ± 0.2 years and 50.7% were female. The majority of participants identified as non-Hispanic White (66.5%), had at least some college education (60.1%), had an annual household income of < $75,000 (65.1%), were current drinkers (65.5%), never smoked or quit smoking for more than one year (79.7%), and reported ‘very good’ or ‘excellent’ general health (60.8%). Approximately 29.5% reported short sleep duration and 19.4% reported having insomnia symptoms (Table 1).

**Table 1.**
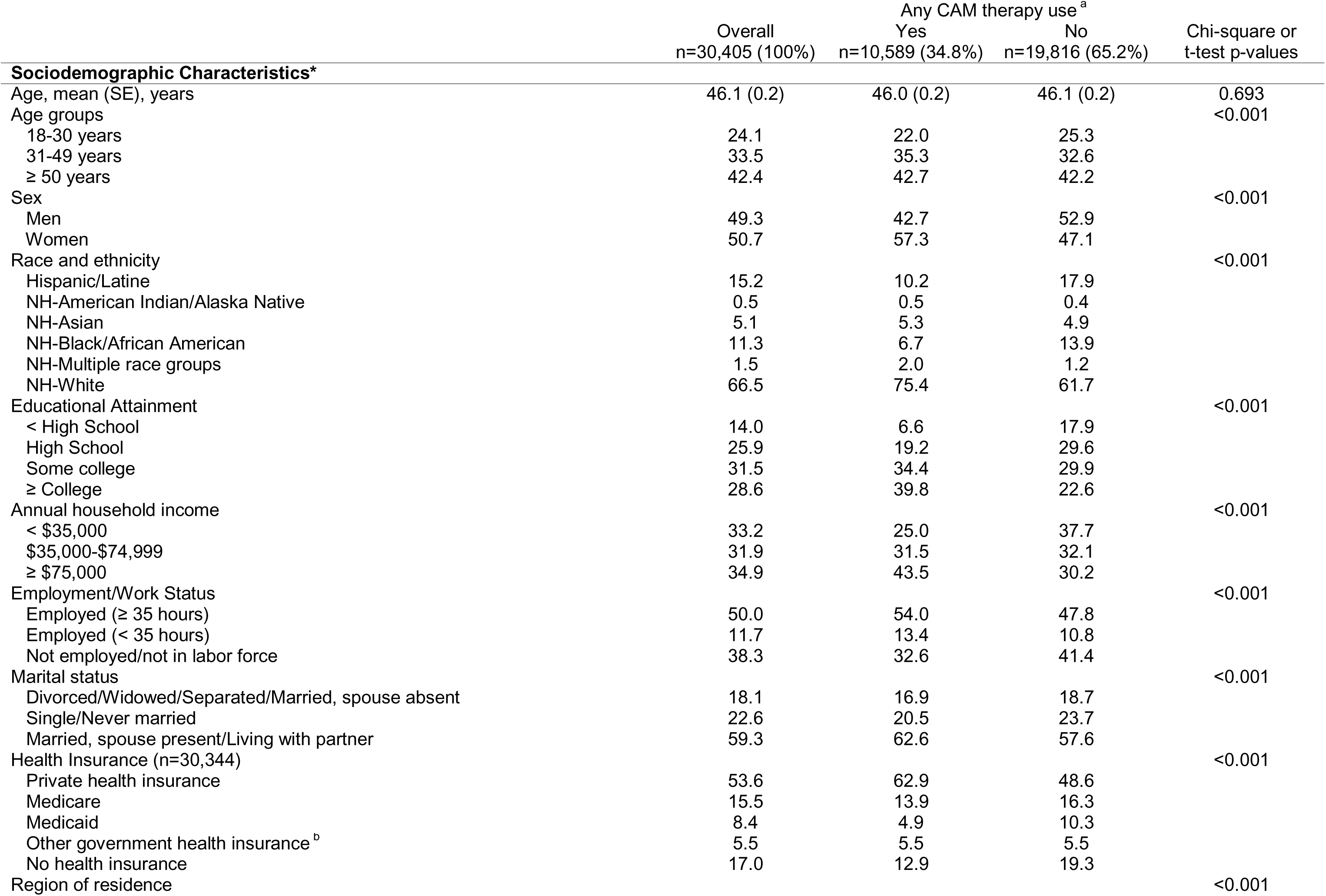
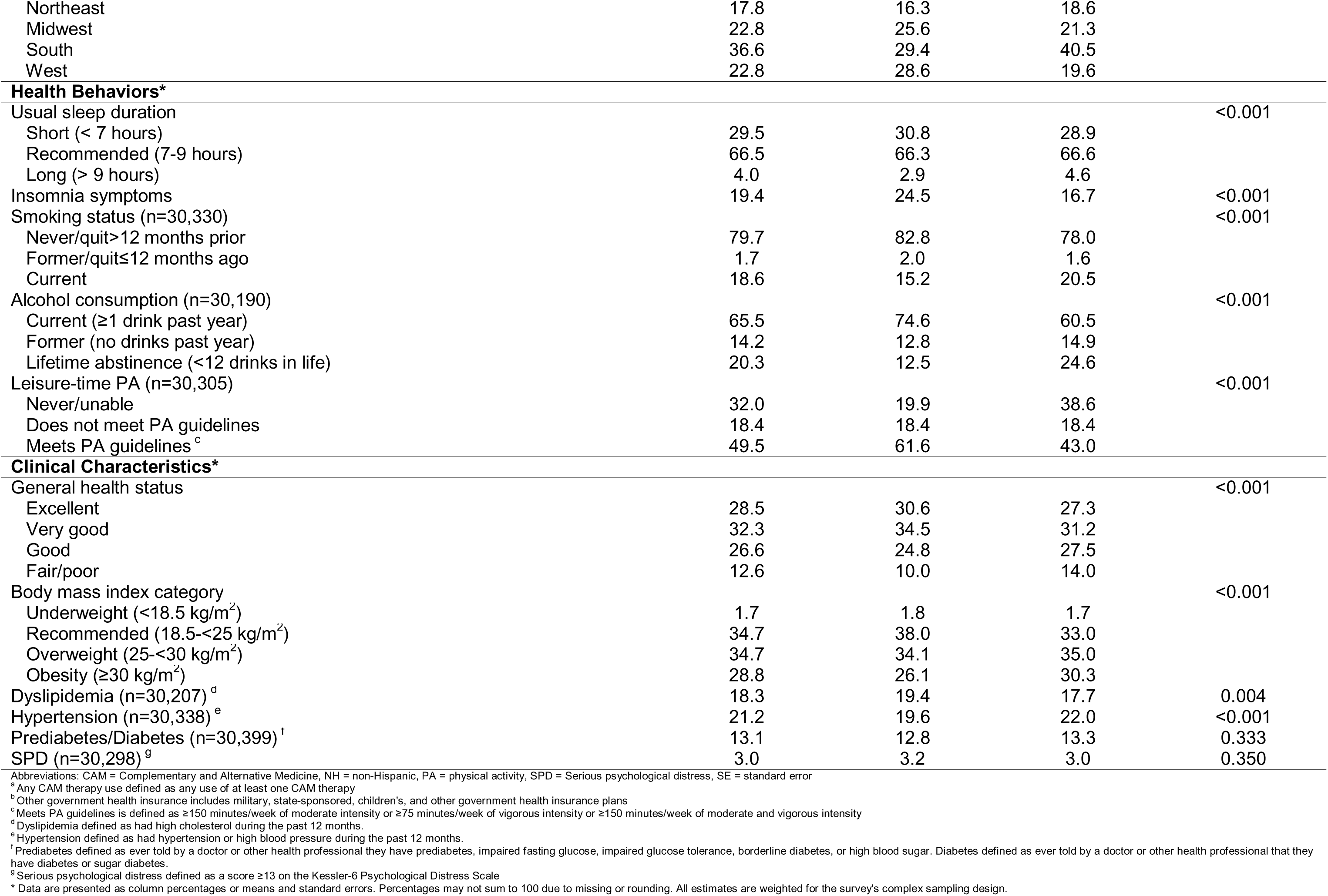
Study population characteristics of adults, overall and stratified by any Complementary and Alternative Medicine use, National Health Interview Survey, 2012, (N=30,405)

Overall, the weighted prevalence of reported CAM therapy use was 35.2%. The mean age was similar between CAM users (46.0 ± 0.2 years) and non-users (46.1 ± 0.2 years). Compared to non-use, CAM utilization was higher among women, NH-White participants, individuals with at least a college degree, participants with an annual household income of ≥$75,000, and employed participants. In contrast, men, NH-Black/African American and Hispanic/Latine participants, individuals with a high school degree or less, participants who report an annual household income of <$35,000, and individuals not employed or not in the labor force were less likely to use CAM therapies. Additionally, short sleep duration was more common among non-users (30.8% vs. 28.9%) insomnia symptoms were more common among CAM users (24.5% vs. 16.7%), while (Table 1).

### Weighted prevalence of Complementary and Alternative Medicine therapies

Overall, herbal supplements (18.2%); yoga, Tai Chi, and Qigong (10.1%); chiropractic or osteopathic manipulation (9.2%); and massage therapy (8.9%) were the most prevalent CAM therapies, while chelation (<0.1%), Ayurveda therapies (0.2%), craniosacral therapy (0.3%), biofeedback (0.3%), and hypnosis (0.3%) were the least prevalent (Table 2). Consistent findings were observed across sleep duration groups. Specifically, herbal supplements remained the most prevalent CAM therapy regardless of sleep duration. Among participants with recommended (7 – 9 hours) or short (< 7) sleep duration, yoga, tai chi, and qigong, and chiropractic or osteopathic manipulation remained the most prevalent therapies, although massage therapy was the second most prevalent therapy among participants with short sleep duration (9.9%). Similar findings were observed across insomnia groups, with massage therapy as the second most prevalent CAM therapy among participants with insomnia symptoms.

**Table 2.** Weighted Prevalence of Complementary and Alternative Medicine Therapies, Overall and stratified by usual sleep duration and insomnia, National Health Interview Survey, 2012, (N=30,405)

|  | Overall<br>n=30,405<br>(100%) | Sleep duration |  |  | Insomnia symptoms |  |
| --- | --- | --- | --- | --- | --- | --- |
|  |  | <7 hours<br>n=9,253<br>(30.4%) | 7-9 hours<br>n=19,916<br>(65.5%) | >9 hours<br>n=1,236<br>(4.1%) | Yes<br>n=6,280<br>(20.7%) | No<br>n=24,125<br>(79.3%) |
| CAM Therapies* |  |  |  |  |  |  |
| At least one CAM therapy <sup>a, b</sup> | 35.2 | 36.6 | 35.1 | 25.6 | 44.4 | 32.9 |
| Acupuncture <sup>b</sup> | 1.6 | 1.4 | 1.7 | 0.9 | 2.3 | 1.4 |
| Ayurveda <sup>b</sup> | 0.2 | 0.1 | 0.3 | 0.3 | 0.4 | 0.2 |
| Biofeedback <sup>b</sup> | 0.3 | 0.3 | 0.3 | 0.7 | 0.5 | 0.3 |
| Chelation Therapy | <0.1 | <0.1 | <0.1 | 0.0 | 0.1 | <0.1 |
| Chiropractic or Osteopathic Manipulation <sup>a, b</sup> | 9.2 | 8.9 | 9.5 | 6.5 | 11.5 | 8.6 |
| Craniosacral Therapy <sup>b</sup> | 0.3 | 0.2 | 0.4 | <0.1 | 0.5 | 0.3 |
| Energy Healing Therapy <sup>b</sup> | 0.7 | 0.7 | 0.7 | 0.7 | 1.4 | 0.5 |
| Herbal Supplement <sup>a, b</sup> | 18.2 | 19.5 | 17.8 | 14.3 | 24.7 | 16.6 |
| Homeopathy <sup>b</sup> | 2.2 | 2.4 | 2.2 | 1.3 | 3.5 | 1.9 |
| Hypnosis <sup>b</sup> | 0.3 | 0.3 | 0.3 | 0.3 | 0.5 | 0.2 |
| Massage <sup>a, b</sup> | 8.9 | 9.9 | 8.7 | 5.0 | 12.4 | 8.1 |
| Meditation, guided imagery, or progressive relaxation <sup>b</sup> | 4.8 | 5.1 | 4.8 | 3.5 | 9.0 | 3.8 |
| Movement or exercise techniques <sup>a</sup> | 2.2 | 1.9 | 2.3 | 1.0 | 2.5 | 2.1 |
| Naturopathy <sup>b</sup> | 0.7 | 0.6 | 0.7 | 0.3 | 1.2 | 0.6 |
| Special diets <sup>b</sup> | 3.1 | 3.0 | 3.1 | 3.0 | 5.1 | 2.6 |
| Traditional Healers <sup>b</sup> | 0.4 | 0.5 | 0.4 | 0.4 | 0.7 | 0.4 |
| Yoga, Tai Chi, or Qigong <sup>a, b</sup> | 10.1 | 9.7 | 10.7 | 4.6 | 11.6 | 9.8 |

Abbreviations: CAM = Complementary and Alternative Medicine
<sup>a</sup> Significant association between CAM therapy and usual sleep duration using Pearson's Chi-Square test (p < 0.05)
<sup>b</sup> Significant association between CAM therapy and insomnia using Pearson's Chi-Square test (p < 0.05)
\* Data are presented as column percentages or means and standard errors. Percentages may not sum to 100 due to missing or rounding. All estimates are weighted for the survey's complex sampling design.

### Complementary and Alternative Medicine therapies and short sleep duration

Participants who reported using at least one type of CAM therapy had a 11% (aPR: 1.11 [1.05 - 1.16]) higher prevalence of short sleep duration, after adjustment, compared to those who did not report using any CAM therapy (Table 3). For individual CAM therapies, herbal supplements (aPR: 1.13 [1.07 - 1.19]) and massage (aPR: 1.14 [1.05 - 1.23]) were associated with a 14% higher prevalence of short sleep duration compared with no CAM use.

**Table 3.**
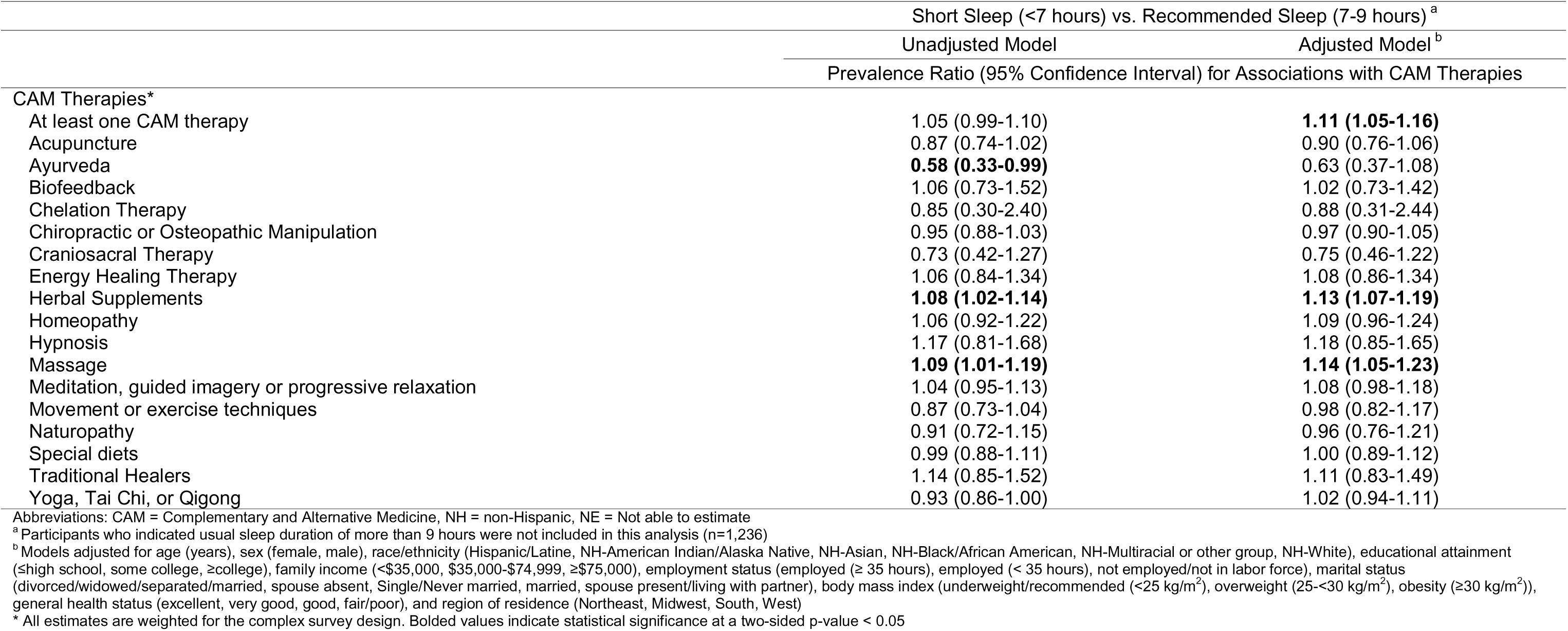
Adjusted and Unadjusted Prevalence Ratios of Short Sleep Duration Associated with Complementary and Alternative Medicine Therapies, National Health Interview Survey, 2012, (N=29,169)

|  | Short Sleep (<7 hours) vs. Recommended Sleep (7-9 hours) <sup>a</sup> |  |
| --- | --- | --- |
|  | Unadjusted Model | Adjusted Model <sup>b</sup> |
|  | Prevalence Ratio (95% Confidence Interval) for Associations with CAM Therapies |  |
| CAM Therapies* |  |  |
| At least one CAM therapy | 1.05 (0.99-1.10) | **1.11 (1.05-1.16)** |
| Acupuncture | 0.87 (0.74-1.02) | 0.90 (0.76-1.06) |
| Ayurveda | **0.58 (0.33-0.99)** | 0.63 (0.37-1.08) |
| Biofeedback | 1.06 (0.73-1.52) | 1.02 (0.73-1.42) |
| Chelation Therapy | 0.85 (0.30-2.40) | 0.88 (0.31-2.44) |
| Chiropractic or Osteopathic Manipulation | 0.95 (0.88-1.03) | 0.97 (0.90-1.05) |
| Craniosacral Therapy | 0.73 (0.42-1.27) | 0.75 (0.46-1.22) |
| Energy Healing Therapy | 1.06 (0.84-1.34) | 1.08 (0.86-1.34) |
| Herbal Supplements | **1.08 (1.02-1.14)** | **1.13 (1.07-1.19)** |
| Homeopathy | 1.06 (0.92-1.22) | 1.09 (0.96-1.24) |
| Hypnosis | 1.17 (0.81-1.68) | 1.18 (0.85-1.65) |
| Massage | **1.09 (1.01-1.19)** | **1.14 (1.05-1.23)** |
| Meditation, guided imagery or progressive relaxation | 1.04 (0.95-1.13) | 1.08 (0.98-1.18) |
| Movement or exercise techniques | 0.87 (0.73-1.04) | 0.98 (0.82-1.17) |
| Naturopathy | 0.91 (0.72-1.15) | 0.96 (0.76-1.21) |
| Special diets | 0.99 (0.88-1.11) | 1.00 (0.89-1.12) |
| Traditional Healers | 1.14 (0.85-1.52) | 1.11 (0.83-1.49) |
| Yoga, Tai Chi, or Qigong | 0.93 (0.86-1.00) | 1.02 (0.94-1.11) |

Abbreviations: CAM = Complementary and Alternative Medicine, NH = non-Hispanic, NE = Not able to estimate
<sup>a</sup> Participants who indicated usual sleep duration of more than 9 hours were not included in this analysis (n=1,236)
<sup>b</sup> Models adjusted for age (years), sex (female, male), race/ethnicity (Hispanic/Latine, NH-American Indian/Alaska Native, NH-Asian, NH-Black/African American, NH-Multiracial or other group, NH-White), educational attainment (≤high school, some college, ≥college), family income (<\$35,000, \$35,000-\$74,999, ≥\$75,000), employment status (employed (≥ 35 hours), employed (< 35 hours), not employed/not in labor force), marital status (divorced/widowed/separated/married, spouse absent, Single/Never married, married, spouse present/living with partner), body mass index (underweight/recommended (<25 kg/m<sup>2</sup>), overweight (25-<30 kg/m<sup>2</sup>), obesity (≥30 kg/m<sup>2</sup>)), general health status (excellent, very good, good, fair/poor), and region of residence (Northeast, Midwest, South, West)
\* All estimates are weighted for the complex survey design. Bolded values indicate statistical significance at a two-sided p-value < 0.05

### Complementary and Alternative Medicine therapies and insomnia symptoms

Overall, the prevalence of insomnia symptoms was 56% higher (aPR: 1.56 [1.47 - 1.65]) among participants who reported using at least one type of CAM therapy compared to those who did not report any CAM therapy use (Table 4). Notably, most CAM therapies, except biofeedback and chelation, were associated with a higher prevalence of insomnia symptoms, with the strongest associations observed for meditation, guided imagery, or progressive relaxation (aPR: 1.84 [1.68 - 2.02]). Point estimates were also high for associations between Ayurveda therapies (aPR: 1.73 [1.17 – 2.57]) and naturopathy (aPR: 1.69 [1.32 – 2.17]) with insomnia symptoms; however, confidence intervals overlapped with those of other individual CAM therapies.

**Table 4.** Adjusted and Unadjusted Prevalence Ratios of Insomnia Associated with Complementary and Alternative Medicine Therapies, National Health Interview Survey, 2012, (N=30,405)

|  | Insomnia Symptoms (Yes vs No) |  |
| --- | --- | --- |
|  | Unadjusted Model | Adjusted Model <sup>a</sup> |
|  | Prevalence Ratio (95% Confidence Interval) for Associations with CAM Therapies |  |
| CAM Therapies* |  |  |
| At least one CAM therapy | **1.47 (1.39-1.56)** | **1.56 (1.47-1.65)** |
| Acupuncture | **1.51 (1.28-1.77)** | **1.44 (1.25-1.66)** |
| Ayurveda | **1.55 (1.04-2.33)** | **1.73 (1.17-2.57)** |
| Biofeedback | **1.75 (1.28-2.41)** | 1.22 (0.92-1.63) |
| Chelation Therapy | 1.47 (0.56-3.88) | 1.62 (0.58-4.51) |
| Chiropractic or Osteopathic Manipulation | **1.28 (1.17-1.41)** | **1.31 (1.20-1.43)** |
| Craniosacral Therapy | **1.69 (1.18-2.42)** | **1.54 (1.13-2.10)** |
| Energy Healing Therapy | **2.00 (1.62-2.48)** | **1.65 (1.33-2.04)** |
| Herbal Supplements | **1.47 (1.37-1.58)** | **1.49 (1.39-1.60)** |
| Homeopathy | **1.58 (1.36-1.84)** | **1.47 (1.25-1.74)** |
| Hypnosis | **1.72 (1.20-2.47)** | **1.59 (1.08-2.36)** |
| Massage | **1.45 (1.34-1.57)** | **1.54 (1.42-1.67)** |
| Meditation, guided imagery or progressive relaxation | **1.94 (1.77-2.13)** | **1.84 (1.68-2.02)** |
| Movement or exercise techniques | 1.15 (0.96-1.39) | **1.35 (1.12-1.63)** |
| Naturopathy | **1.71 (1.32-2.22)** | **1.69 (1.32-2.17)** |
| Special diets | **1.71 (1.51-1.93)** | **1.62 (1.42-1.85)** |
| Traditional Healers | **1.63 (1.24-2.13)** | **1.53 (1.16-2.01)** |
| Yoga, Tai Chi, or Qigong | **1.16 (1.06-1.28)** | **1.33 (1.21-1.46)** |

Abbreviations: CAM = Complementary and Alternative Medicine, NH = non-Hispanic, NE = Not able to estimate
<sup>a</sup> Models adjusted for age (years), sex (female, male), race/ethnicity (Hispanic/Latine, NH-American Indian/Alaska Native, NH-Asian, NH-Black/African American, NH-Multiracial or other group, NH-White), educational attainment (≤high school, some college, ≥college), family income (<\$35,000, \$35,000-\$74,999, ≥\$75,000), employment status (employed (≥ 35 hours), employed (< 35 hours), not employed/not in labor force), marital status (divorced/widowed/separated/married, spouse absent, Single/Never married, married, spouse present/living with partner), body mass index (underweight/recommended (<25 kg/m<sup>2</sup>), overweight (25-30 kg/m<sup>2</sup>), obesity (≥30 kg/m<sup>2</sup>)), general health status (excellent, very good, good, fair/poor), and region of residence (Northeast, Midwest, South, West)
\* All estimates are weighted for the complex survey design. Bolded values indicate statistical significance at a two-sided p-value < 0.05

## DISCUSSION

In this nationally representative sample of US adults, CAM therapy use was associated with a higher prevalence of both short sleep duration and insomnia symptoms. Among individual CAM therapies, herbal supplements and massage were associated with a higher prevalence of short sleep duration. Most CAM therapies examined were associated with a higher prevalence of insomnia symptoms, and the strongest associations were observed for meditation/guided imagery/progressive relaxation with suggestion of strong associations for naturopathy and Ayurveda therapies. These findings highlight the widespread use of CAM therapies among adults with sleep-related concerns and suggest that individuals experiencing poor sleep may be more likely to seek CAM approaches as part of sleep management strategies.

Our findings are generally consistent with prior studies demonstrating high CAM utilization among individuals with insomnia and sleep disturbances [17, 30, 42]. Prior national survey studies have shown that adults reporting insomnia symptoms are significantly more likely to use relaxation techniques, mind-body therapies, herbal supplements, and other CAM approaches compared to adults without sleep difficulties [17, 42]. Existing literature has primarily focused on whether specific CAM interventions improve self-reported sleep quality, insomnia severity, sleep latency, or related symptoms, particularly mindfulness-based movement and mind-body therapies such as yoga, Tai Chi, Qigong, meditation, acupuncture, acupressure, and relaxation techniques [30, 36, 37, 43, 44]. Recent umbrella review and systematic reviews suggest that several CAM approaches may improve subjective sleep outcomes among individuals with insomnia, although findings remain heterogeneous and evidence quality is often limited by small sample sizes, methodological variability, and reliance on self-reported sleep measures [36–38, 43].

Our study builds upon prior research by illuminating more recent nationally-representative associations – albeit cross sectional – between a broad range of CAM therapies and both short sleep duration and insomnia symptoms among a sample of US adults. Importantly, the observed associations should not be interpreted as evidence that CAM therapies worsen sleep. Rather, these findings are consistent with possible reverse causation, confounding by indication, or symptom-driven healthcare-seeking behaviors, whereby individuals experiencing chronic insomnia symptoms, insufficient sleep, stress, anxiety, chronic pain, or other health concerns may be more likely to pursue CAM therapies as an attempt (even if experimental) to alleviate symptoms in hopes of improving sleep and overall well-being [17, 42]. Prior studies similarly suggest that many adults use CAM approaches not solely to treat insomnia itself, but also to manage co-occurring psychological distress, stress-related symptoms, chronic medical conditions, and perceived declines in quality of life [17, 42]. Because insomnia frequently co-occurs with cardiometabolic disease, anxiety, depression, chronic pain, and psychosocial stressors, individuals experiencing more severe or persistent sleep difficulties may be particularly motivated to seek holistic, non-pharmacologic, or self-directed approaches for symptom management [37, 38]. This may also explain why associations were observed not only for insomnia symptoms, but also for short sleep duration, as insufficient sleep may represent a marker of, for example, broader psychological distress, occupational demands, caregiving burden, chronic disease burden, or stress-related sleep disruption that motivates CAM utilization.

The strong associations observed between meditation, guided imagery, or progressive relaxation and insomnia symptoms likely reflect both treatment-seeking behavior among individuals with persistent sleep problems and the established role of these therapies as nonpharmacologic approaches for managing insomnia. Prior evidence, including systematic reviews and network meta-analysis of randomized controlled trials, supports the effectiveness of mindfulness-and relaxation-based therapies for improving sleep quality and reducing insomnia symptoms [30, 36–38, 43]. However, given that our study was cross-sectional, these associations may also reflect that individuals with more severe, chronic, or treatment-resistant insomnia are especially likely to pursue mind-body therapies. Prior studies suggest that many adults seek CAM therapies as part of self-management of sleep problems, often because of dissatisfaction with conventional treatments, concerns regarding medication side effects or dependency, limited access to cognitive behavioral therapy for insomnia, or preferences for more holistic and person-centered approaches to care [37, 38, 43]. Similarly, the possible stronger associations of naturopathy and Ayurveda therapies with insomnia symptoms (than associations with other therapies) may reflect higher chronic symptom burden or preferences for culturally congruent and integrative approaches to health and healing. Ayurveda and other traditional healing systems conceptualize sleep as interconnected with spiritual, emotional, behavioral, dietary, and environmental balance, emphasizing individualized, integrative, and whole-person approaches to care [38]. Likewise, Traditional Chinese Medicine, meditation, yoga-based practices, and herbal therapies are often rooted in broader philosophies emphasizing mind-body integration, restoration of balance, stress reduction, and preventive health behaviors [37, 45]. Such approaches may be particularly appealing among individuals seeking alternatives or complements to pharmacologic sleep treatments or among individuals whose cultural beliefs and health practices align more closely with integrative or traditional systems of medicine.

The higher prevalence of CAM use among females, individuals with higher educational attainment, and those with higher household income is also consistent with prior literature describing the sociodemographic profile of CAM users in the United States [17, 24, 46]. Prior studies suggest that women are generally more proactive than men in healthcare utilization and may be more open to mind-body and integrative approaches emphasizing self-care, stress management, and wellness promotion [46]. Additionally, women experience a disproportionately higher prevalence of insomnia, sleep disturbances, anxiety, caregiving burden, and stress-related conditions across the lifespan, which may further contribute to greater CAM utilization [47]. Higher educational attainment and income may additionally facilitate awareness of CAM therapies, access to integrative health services, and the financial ability to afford therapies that are often not covered by insurance and require substantial out-of-pocket costs [46]. In contrast, lower reported CAM utilization among non-Hispanic Black/African American and Hispanic/Latine participants may reflect structural barriers to integrative healthcare access, differences in healthcare utilization patterns and health beliefs, financial constraints, and potential underrepresentation of culturally-rooted healing practices within standardized national CAM survey measures [48]. More research is warranted.

Several limitations of this study should be considered. First, the cross-sectional design precludes causal inference and limits the ability to determine the temporal direction of associations between CAM use and sleep outcomes. As such, findings may reflect reverse causation or confounding by indication, whereby adults with more severe sleep disturbances are more likely to seek CAM therapies. Second, sleep duration, insomnia symptoms, and CAM use were self-reported and therefore subject to recall bias and reporting bias, resulting in possible misclassification. Third, the NHIS did not capture important contextual information regarding frequency, duration, timing, reasons for CAM use, treatment adherence, or perceived effectiveness, limiting the interpretation of whether CAM therapies improved, worsened, or were unrelated to sleep outcomes over time. Fourth, some CAM modalities had relatively low prevalence estimates, which reduced statistical precision. Additionally, standardized national CAM survey measures did not fully capture culturally rooted healing traditions, informal health practices, or community-based approaches commonly used across diverse populations, potentially underestimating utilization of certain integrative or traditional practices. Finally, residual confounding from unmeasured psychosocial, behavioral, occupational, or health-related factors likely remains.

Despite the limitations, our study also has important strengths. First, the analyses were conducted using a large, nationally representative sample of US adults, enhancing the generalizability of findings and allowing examination of multiple CAM therapies across diverse sociodemographic groups. Second, unlike many prior studies focused primarily on intervention efficacy within clinical samples, this study examined population-level associations between CAM utilization and both short sleep duration and insomnia symptoms, providing broader insight into patterns of CAM use among adults experiencing sleep difficulties. Additionally, the inclusion of multiple CAM modalities, including herbal supplements, meditation and relaxation techniques, acupuncture, naturopathy, and Ayurveda therapies, allowed for a more comprehensive evaluation of associations across a broad range of integrative and traditional health practices. The study also contributes to a growing literature examining sleep health within whole-person and integrative health frameworks.

Our findings have several important clinical and public health implications. Sleep disturbances are common, burdensome, and frequently co-occur with stress, chronic pain, anxiety, depression, and cardiometabolic conditions, leading many individuals to seek holistic and non-pharmacologic approaches to symptom management. The high prevalence of CAM use among adults reporting insomnia symptoms and short sleep duration highlights the growing role of integrative and mind-body approaches in sleep health management and underscores the importance of clinicians routinely assessing CAM use during clinical encounters. Increased awareness of CAM utilization may facilitate more patient-centered discussions regarding treatment preferences, potential benefits and risks, herb-drug interactions, and integration of evidence-based non-pharmacologic therapies into sleep care. These findings also support growing interest in whole-person and integrative approaches to sleep health that consider behavioral, psychological, spiritual, social, cultural, and environmental contributors to sleep disturbances [27, 29, 37]. Mind-body therapies such as meditation, yoga, Tai Chi, and relaxation-based interventions may hold particular promise given their potential to simultaneously target stress, emotional well-being, hyperarousal, and sleep-related symptoms [36]. However, additional longitudinal studies and rigorous clinical trials are needed to clarify the effectiveness of specific CAM therapies for insomnia symptoms and insufficient sleep across diverse populations and clinical contexts. Future research should also investigate motivations for CAM use, barriers to accessing evidence-based sleep care, and how culturally congruent and integrative approaches may support sleep health promotion and chronic disease prevention.

In conclusion, CAM therapy use was common and associated with a higher prevalence of both short sleep duration and insomnia symptoms in this nationally representative sample of US adults. Mind-body therapies, herbal supplements, and naturopathy therapies demonstrated particularly strong, positive associations with sleep-related difficulties. These findings likely reflect increased utilization of CAM therapies among individuals experiencing chronic or burdensome sleep disturbances rather than harmful effects of CAM therapies themselves. As public interest in integrative, holistic, and non-pharmacologic approaches to health continues to grow, greater attention to CAM use within sleep medicine and public health research more generally may help inform more patient-centered and culturally-responsive approaches to sleep health promotion. Future longitudinal and intervention studies are needed to better understand the temporal relationship between CAM use and sleep outcomes and to identify which therapies may be most effective for improving sleep health and overall well-being.

## DECLARATIONS

### Ethics approval and consent to participate

Not applicable

### Consent for publication

All authors have seen and approved the manuscript.

### Availability of data and materials

The datasets analyzed during the current study are publicly available from the National Center for Health Statistics (NCHS), Centers for Disease Control and Prevention, National Health Interview Survey (NHIS), at: https://www.cdc.gov/nchs/nhis/index.htm

## Data Availability

All data produced in the present study are available upon reasonable request to the authors

## Acknowledgements

The authors would like to thank all respondents who participated in the NHIS survey.

## Author contributions

*Authors:* Rupsha Singh, Symielle A. Gaston, Christopher Payne, Dayna T. Neo, Suzanne M. Bertisch, Chandra L. Jackson

*Study concept:* Chandra L. Jackson.

*Study design:* Rupsha Singh, Symielle A. Gaston, Christopher Payne, Chandra L. Jackson.

*Acquisition of data:* Christopher Payne, CL. Jackson.

*Statistical Analysis:* Christopher Payne.

*Interpretation of data:* Rupsha Singh, Symielle A. Gaston, Christopher Payne, Dayna T. Neo, Suzanne M. Bertisch, Chandra L. Jackson.

*Drafting of the manuscript:* Rupsha Singh, Christopher Payne, Dayna T. Neo.

*Critical revision of the manuscript for important intellectual content:* Rupsha Singh, Symielle A. Gaston, Christopher Payne, Dayna T. Neo, Suzanne M. Bertisch, Chandra L. Jackson.

*Administrative, technical, and material support:* CL. Jackson.

*Obtaining funding and study supervision:* CL. Jackson.

*Final Approval:* Rupsha Singh, Symielle A. Gaston, Christopher Payne, Dayna T. Neo, Suzanne M. Bertisch, Chandra L. Jackson.

---

**Supplemental Figure 1.**
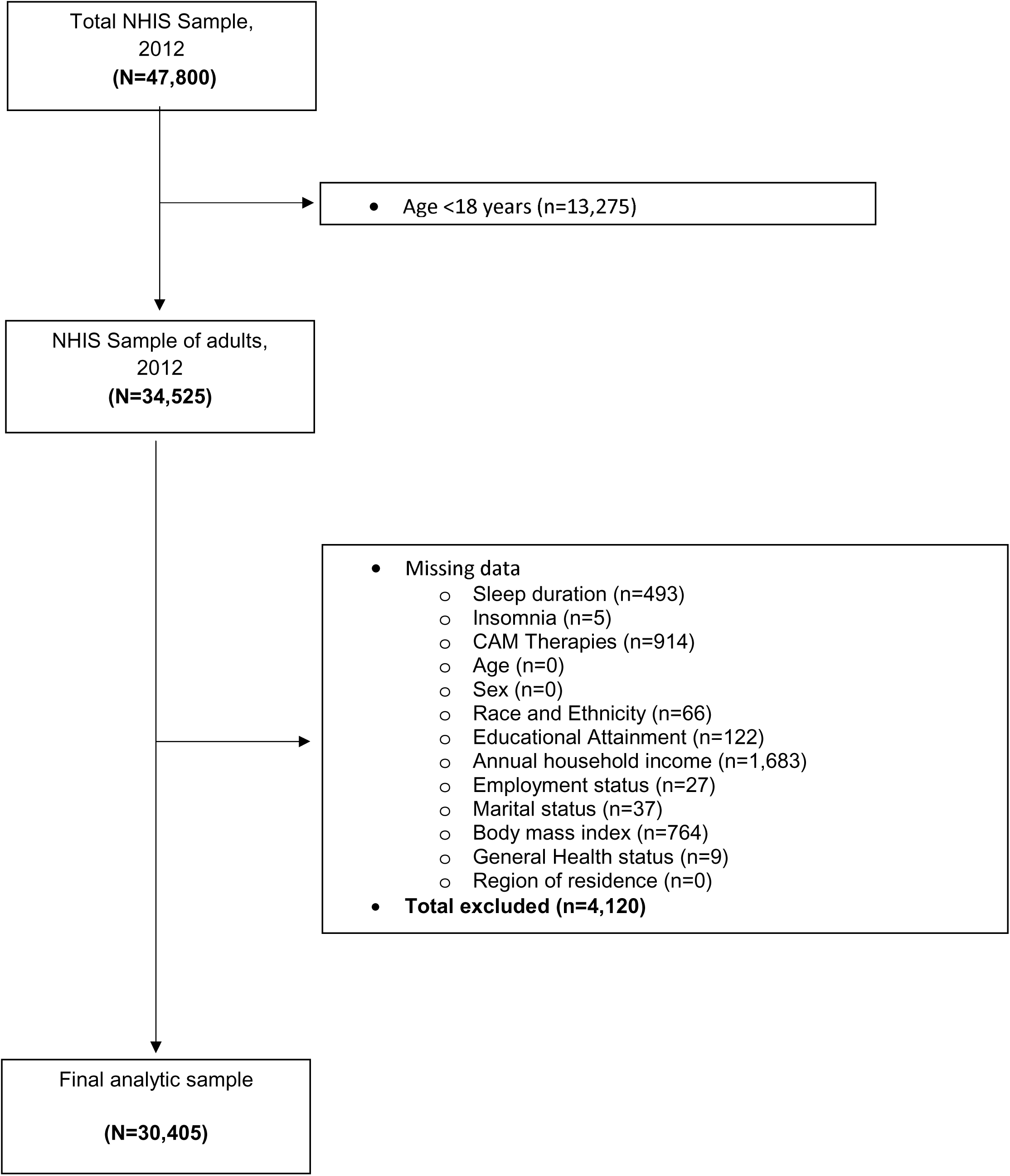
Flow chart of study population selection

**Supplemental Table 1.** Comparison of included and excluded study participants, National Health Interview Survey, 2012, (N=34,525)

|  | Included<br>n=30,405 (88.1%) | Excluded<br>n=4,120 (11.9%) | Chi-square or<br>t-test p-value |
| --- | --- | --- | --- |
| **Sociodemographic Characteristics\*** |  |  |  |
| Age (years), mean (SE) | 46.1 (0.2) | 50.4 (0.4) | <0.001 |
| Age group |  |  | <0.001 |
| 18-30 years | 24.1 | 18.3 |  |
| 31-49 years | 33.5 | 29.2 |  |
| ≥ 50 years | 42.4 | 52.5 |  |
| Sex |  |  | <0.001 |
| Men | 49.3 | 39.7 |  |
| Women | 50.7 | 60.3 |  |
| Race/ethnicity |  |  | 0.002 |
| Hispanic/Latine | 15.2 | 12.9 |  |
| NH-American Indian/Native Alaskan | 0.5 | 0.8 |  |
| NH-Asian | 5.1 | 5.8 |  |
| NH-Black/African American | 11.3 | 12.5 |  |
| NH-Multiple race groups | 1.5 | 0.9 |  |
| NH-White | 66.5 | 67.0 |  |
| Educational Attainment |  |  | 0.003 |
| <High School | 14.0 | 14.3 |  |
| High School graduate | 25.9 | 29.3 |  |
| Some College | 31.5 | 30.0 |  |
| ≥College | 28.6 | 26.4 |  |
| Annual household income |  |  | <0.001 |
| < \$35,000 | 33.2 | 39.2 | |
| \$35,000-\$74,999 | 31.9 | 32.6 | |
| ≥ \$75,000 | 34.9 | 28.2 | |
| Employment/Work status |  |  | <0.001 |
| Employed (≥ 35 hours) | 50.0 | 42.8 |  |
| Employed (< 35 hours) | 11.7 | 11.2 |  |
| Not employed/not in labor force | 38.3 | 46.0 |  |
| Marital status |  |  | 0.002 |
| Divorced/Widowed/Separated/Married, spouse absent | 18.1 | 20.7 |  |
| Single/Never married | 22.6 | 20.6 |  |
| Married, spouse present/living with partner | 59.3 | 58.7 |  |
| Health insurance (n=34,413) |  |  | <0.001 |
| Private health insurance | 53.6 | 47.3 |  |
| Medicare | 15.5 | 22.4 |  |
| Medicaid | 8.4 | 8.4 |  |
| Other government health insurance<sup>a</sup> | 5.5 | 5.6 |  |
| No health insurance | 17.0 | 16.4 |  |
| Region of residence |  |  | 0.014 |
| Northeast | 17.8 | 20.9 |  |
| Midwest | 22.8 | 22.0 |  |
| South | 36.6 | 35.2 |  |
| West | 22.8 | 21.9 |  |
| **Health Behaviors\*** |  |  |  |
| Usual sleep duration |  |  | 0.002 |
| Short (<7 hours) | 29.5 | 26.3 |  |
| Recommended (7-9 hours) | 66.5 | 69.2 |  |
| Long (>9 hours) | 4.0 | 4.6 |  |
| Insomnia symptoms | 19.4 | 18.0 | 0.082 |
| Smoking status (n=34,181) |  |  | <0.001 |
| Never/quit >12 months prior | 79.7 | 83.9 |  |
| Former/quit ≤12 months ago | 1.7 | 1.7 |  |
| Current | 18.6 | 14.4 |  |
| Alcohol consumption (n=33,900) |  |  | <0.001 |
| Current (≥1 drink past year) | 65.5 | 57.3 |  |
| Former (no drinks past year) | 14.2 | 16.6 |  |
| Lifetime abstinence (<12 drinks in life) | 20.3 | 26.1 |  |
| Leisure-time PA (n=34,143) |  |  | <0.001 |
| Never/unable | 32.0 | 38.5 |  |
| Does not meet PA guidelines | 18.4 | 17.3 |  |
| Meets PA guidelines<sup>b</sup> | 49.5 | 44.2 |  |
| Clinical Characteristics* |  |  |  |
| General health status |  |  | <0.001 |
| Excellent | 28.5 | 26.2 |  |
| Very good | 32.3 | 30.9 |  |
| Good | 26.6 | 27.7 |  |
| Fair/poor | 12.6 | 15.3 |  |
| Body mass index category |  |  | <0.001 |
| Underweight (<18.5 kg/m<sup>2</sup>) | 1.7 | 1.9 |  |
| Recommended (18.5-<25 kg/m<sup>2</sup>) | 34.7 | 38.0 |  |
| Overweight (25-<30 kg/m<sup>2</sup>) | 34.7 | 36.0 |  |
| Obesity (≥30 kg/m<sup>2</sup>) | 28.8 | 24.1 |  |
| Dyslipidemia (n=34,264)<sup>c</sup> | 18.3 | 18.7 | 0.596 |
| Hypertension (n=34,420)<sup>d</sup> | 21.2 | 24.1 | 0.001 |
| Prediabetes/Diabetes (n=34,506)<sup>e</sup> | 13.1 | 14.5 | 0.076 |
| SPD (n=34,306)<sup>f</sup> | 3.0 | 2.7 | 0.348 |

Abbreviations: CAM = Complementary and Alternative Medicine, NH = non-Hispanic, PA = physical activity, SPD = Serious psychological distress, SE = standard error
<sup>a</sup> Other government health insurance includes military, state-sponsored, children's, or other government health insurance plans
<sup>b</sup> Meets PA guidelines is defined as ≥150 minutes/week of moderate intensity or ≥75 minutes/week of vigorous intensity or ≥150 minutes/week of moderate and vigorous intensity
<sup>c</sup> Dyslipidemia defined as had high cholesterol during the past 12 months.
<sup>d</sup> Hypertension defined as had hypertension or high blood pressure during the past 12 months.
<sup>e</sup> Prediabetes defined as ever told by a doctor or other health professional they have prediabetes, impaired fasting glucose, impaired glucose tolerance, borderline diabetes, or high blood sugar. Diabetes defined as ever told by a doctor or other health professional that they have diabetes or sugar diabetes.
<sup>f</sup> Serious psychological distress defined as a score ≥13 on the Kessler-6 Psychological Distress Scale
\* Data are presented as column percentages or means and standard errors. Percentages may not sum to 100 due to missing or rounding. All estimates are weighted for the survey's complex sampling design.

## Appendix I – CAM therapies

### Acupuncture

The 2012 field representative’s manual defines acupuncture as a family of procedures involving stimulation of anatomical points on the body by a variety of techniques. American practices of acupuncture incorporate medical traditions from China, Japan, Korea, and other countries. The acupuncture technique most studied scientifically involves penetrating the skin with thin, solid, metallic needles that are manipulated by hand or by electrical stimulation.

### Ayurveda

The 2012 field representative’s manual defines ayurveda as a system of medicine that originated in India several thousand years ago. In the United States, Ayurveda is considered a type of CAM and a whole medical system. As with other such systems, it is based on theories of health and illness and on ways to prevent, manage, or treat health problems. Ayurveda aims to integrate and balance the body, mind, and spirit (thus, some view it as “holistic”). This balance is believed to lead to contentment and health and to help prevent illness. However, Ayurveda also proposes treatments for specific health problems, whether they are physical or mental. An aim of Ayurvedic practices is to cleanse the body of substances that can cause disease, and this is believed to help reestablish harmony and balance.

### Biofeedback

The 2012 field representative’s defines biofeedback as a therapy that uses simple electronic devices to teach clients how to consciously regulate bodily functions, such as breathing, heart rate, and blood pressure, to improve overall health. Biofeedback is used to reduce stress, eliminate headaches, recondition injured muscles, control asthmatic attacks, and relieve pain.

### Chelation therapy

The 2012 field representative’s manual defines chelation therapy as a chemical process in which a substance is used to bind molecules, such as metals or minerals, and hold them tightly so that they can be removed from a system, such as the body. In medicine, chelation has been scientifically proven to rid the body of excess or toxic metals. For example, a person who has lead poisoning may be given chelation therapy to bind and remove excess lead from the body before it can cause damage.

### Chiropractic or osteopathic manipulation

The 2012 field representative’s manual defines chiropractic manipulation as a form of health care that focuses on the relationship between the body’s structure, primarily of the spine, and function. Chiropractors or chiropractic physicians use a type of hands-on therapy called manipulation (or adjustment) as their core clinical procedure.

The 2012 field representative’s manual defines osteopathic manipulation as a full-body system of hands-on techniques to alleviate pain, restore function, and promote health and well-being.

### Craniosacral therapy

The 2012 field representative’s manual defines craniosacral therapy as a body-based practice. Practitioners use light touch and manipulation focused on the skull and spine, with the intent of sensing and removing what they refer to as blockages or imbalances that may be contributing to a health condition.

### Energy healing therapy

The 2012 field representative’s manual defines energy healing therapy as the channeling of healing energy through the hands of a practitioner into the client’s body to restore a normal energy balance and, therefore, health. Energy healing therapy has been used to treat a wide variety of ailments and health problems and is often used in conjunction with other alternative and conventional medical treatments.

### Homeopathy

The 2012 field representative’s manual defines homeopathy as a system of medical practices based on the theory that any substance that can produce symptoms of disease or illness in a healthy person can cure those symptoms in a sick person. For example, someone suffering from insomnia may be given a homeopathic dose of coffee. Administered in diluted form, homeopathic remedies are derived from many natural sources, including plants, animals, metals, and minerals.

### Hypnosis

The 2012 field representative’s manual defines hypnosis as an altered state of consciousness characterized by increased responsiveness to suggestion. This hypnotic state is attained by first relaxing the body, then shifting attention toward a narrow range of objects or ideas as suggested by the hypnotist or hypnotherapist. The procedure is used to effect positive changes and to treat numerous health conditions including ulcers, chronic pain, respiratory ailments, stress, and headaches.

### Massage therapy

The 2012 field representative’s manual defines massage therapy as the manipulation of muscle and connective tissue to enhance function of those tissues and promote relaxation and well-being.

### Naturopathy

The 2012 field representative’s manual defines naturopathy as an alternative medical system. Naturopathic medicine proposes that there is a healing power in the body that establishes, maintains, and restores health. Practitioners work with the patient with a goal of supporting this power through treatments such as nutrition and lifestyle counseling, dietary supplements, medicinal plants, exercise, homeopathy, and treatments from traditional Chinese medicine.

### Yoga/Tai chi/Qigong

The 2012 field representative’s manual defines yoga as a technique that combines breathing exercises, physical postures, and meditation to calm the nervous system and balance body, mind, and spirit. Usually performed in classes, sessions are conducted once a week or more and roughly last 45 minutes.

The 2012 field representative’s manual defines Tai chi as a mind-body practice that originated in China as a martial art. A person doing tai chi moves his body slowly and gently, while breathing deeply and meditating (tai chi is sometimes called “moving meditation”). Many practitioners believe that tai chi helps the flow throughout the body of a proposed vital energy called “qi.” A person practicing tai chi moves her body in a slow, relaxed, and graceful series of movements. One can practice on one’s own or in a group. The movements make up what are called forms (or routines).

The 2012 field representative’s manual defines Qigong as an ancient Chinese discipline combining the use of gentle physical movements, mental focus, and deep breathing directed toward specific parts of the body. Performed in repetitions, the exercises are normally performed two or more times a week for 30 minutes at a time.

## Appendix II – Traditional healers

### Curandero, Machi or Parchero

The 2007 and later field representative’s manuals state that a Curandero, Machi or Parchero are a type of traditional folk healer. Originally found in Latin America, Curanderos specialize in treating illness through the use of supernatural forces, herbal remedies, and other natural medicines. “Machi” most commonly refers to a traditional healer, shaman, and spiritual leader within the Mapuche culture of Chile and Argentina. These individuals are respected figures with deep knowledge of medicinal herbs, dream interpretation, and spiritual rituals.

### Hierbero, Yerbero or Hierbista

The 2007 and later field representative’s manuals define a Hierbo, Yerbera, or (starting in 2012) a Hierbista or Yerbero as a practitioner with knowledge of the medicinal qualities of plants.

### Huesero

The 2012 field representative’s manual states that a Huesero or “bone setter” (in Hispanic folk healing), specializes in bone ailments, mainly lesions and fractures.

### Native American healers/Medicine men

The 2007 and later field representative’s manuals state that Native American healers/Medicine men use information from the “spirit world” to benefit the community. People see Native American healers for a variety of reasons, especially to find relief or a cure from illness or to find spiritual guidance.

### Shaman

The 2007 and later field representative’s manuals state that Shamans are said to act as mediums between the invisible spiritual world and the physical world. Most gain knowledge through contact with the spiritual world and use the information to perform tasks such as divination, influencing natural events, and healing the sick or injured.

### Sobador

The 2007 and later field representative’s manuals state that a Sobador uses massage and rub techniques to treat patients.

## Appendix III – Herbal and non-vitamin supplements

Acai (pills, gelcaps),

Bee Pollen and other Bee products,

Chondroitin,

Co-enzyme Q10 (CoQ10),

Cranberry (pills or capsules),

Digestive Enzymes (lactaid),

Echinacea,

Fish Oil or omega 3 or DHA fatty acid or EPA fatty acid supplements,

Garlic supplements (pills, gelcaps),

Ginkgo Biloba,

Ginseng,

Glucosamine,

Green tea pills (not brewed tea) or EGCG (pills),

Melatonin,

Milk Thistle (silymarin),

MSM (Methylsulfonylmethane),

Probiotics or Prebiotics,

SAM-e,

Saw Palmetto,

Valerian

## Appendix IV – Meditation, guided imagery, and progressive relaxation techniques

The 2012 field representative’s manual defines meditation as a group of techniques, most of which started in eastern religious or spiritual traditions. In meditation, a person learns to focus their attention and suspend the stream of thoughts that normally occupy the mind. This practice is believed to result in a state of greater physical relaxation, mental calmness, and psychological balance. Practicing meditation can change how a person relates to the flow of emotions and thoughts in the mind. In mantra meditation, the meditator focuses on a mantra (a specially chosen word, sound, or phrase repeated silently). Mindfulness meditation is a type of meditation based on the concept of being mindful, or having increased awareness, of the present. It uses breathing methods, guided imagery, and other practices to relax the body and mind and help reduce stress. It is also known as mindfulness relaxation and mindfulness-based stress reduction. Spiritual meditation may be performed according to the practices of one of the major religions or within a spiritual tradition. The techniques used may be the same as in other types of meditation (for example, transcendental meditation), but the focus is on spirituality (such as repeating a spiritual, meditative phrase).

The 2012 field representative’s manual states that guided imagery is used for healing or health maintenance and involves a series of relaxation techniques followed by the visualization of detailed images, usually calm and peaceful in nature. If used for treatment, the individual will visualize their body free of the specific problem or condition. Sessions are typically 20 to 30 minutes long and may be practiced several times a week.

The 2012 field representative’s manual states that progressive relaxation is used to relieve tension and stress by systematically tensing and relaxing successive muscle groups.

## Appendix V – Movement and exercise techniques

### Alexander technique

The 2012 field representative’s manual defines the Alexander technique as a practice that uses guidance and education on ways to improve posture and movement. The intent is to teach a person how to use muscles more efficiently in order to improve the overall functioning of the body. Examples of the Alexander technique as CAM are using it to treat low-back pain and the symptoms of Parkinson’s disease.

### Feldenkrais method

The 2012 field representative’s manual defines the Feldenkrais method as a method of education in physical coordination and movement. Practitioners use verbal guidance and light touch to teach the method through one-on-one lessons and group classes. The intent is to help the person become more aware of how the body moves through space and to improve physical functioning.

### Pilates

The 2012 field representative’s manual defines Pilates as a method of physical exercise used to strengthen and build control of muscles, especially those used for posture. Awareness of breathing and precise control of movements are integral components of Pilates. Special equipment, if available, is often used.

### Trager Psychophysical Integration

The 2012 field representative’s manual defines Trager Psychophysical Integration as a therapy in which practitioners apply a series of gentle, rhythmic rocking movements to the joints. They also teach physical and mental self-care exercises to reinforce the proper movement of the body. The intent is to release physical tension and increase the body’s range of motion. An example of Trager Psychophysical Integration as CAM is using it to treat chronic headaches.

## Appendix VI – Special diets

### The Atkins diet

The 2012 field representative’s manual states that The Atkins diet emphasizes a drastic reduction in the daily intake of carbohydrates (40 grams or less), countered by an increase in protein and fat.

### Macrobiotic diet

The 2012 field representative’s manual states that a macrobiotic diet is low in fat, emphasizes whole grains and vegetables, and restricts the intake of fluids. Of particular importance is the consumption of fresh, non-processed foods.

### The Ornish diet

The 2012 field representative’s manual states that The Ornish diet is a high fiber, low-fat vegetarian diet that promotes weight loss and health by controlling what one eats, not by restricting the intake of calories. Fruits, beans, grains, and vegetables can be eaten at all meals, while non-fat dairy products such as skim milk, non-fat cheeses, and egg whites are to be consumed in moderation. Products such as oils, avocados, nuts and seeds, and meats of all kind are avoided.

### The Pritikin diet (or Pritikin Principle)

The 2012 field representative’s manual states that while meat is allowed, the Pritikin diet (or Pritikin Principle) is low-fat and emphasizes the consumption of foods with a large volume of fiber and water, including many vegetables, fruits, beans, and natural, unprocessed grains.

### Vegetarian diet

The 2012 field representative’s manual defines a vegetarian diet as totally devoid of meat, red or white. There are, however, numerous variations on the non-meat theme. For example, some vegetarian diets are restricted to plant products only, while others may include eggs and dairy products. Another variation limits consumption to raw fruit, sometimes supplemented with nuts and vegetables. Finally, a number of vegetarian diets prohibit alcohol, sugar, caffeine, or processed foods.

